# The Brazilian COVID-19 vaccination campaign: A modelling analysis of socio-demographic factors on uptake

**DOI:** 10.1101/2023.04.21.23288730

**Authors:** Sabrina L. Li, Carlos A. Prete, Alexander E. Zarebski, Andreza A. de Souza Santos, Ester C. Sabino, Vitor H. Nascimento, Chieh-Hsi Wu, Jane P. Messina

## Abstract

The COVID-19 pandemic has caused over half a million deaths in Brazil, and public healthcare nearly collapsed. Vaccination differs between states and demographics. Dose shortages delayed access.

In this cross-sectional study, data were retrieved from the Brazilian Ministry of Health databases published since 17 January 2021, respectively. We developed a campaign optimality index to characterise inequality in vaccination access caused by age due to premature vaccination towards younger populations before older and vulnerable populations were fully vaccinated. We assessed geographical inequalities in full vaccination coverage and dose by age, sex, race, and socioeconomic status. Generalised linear regression was used to investigate the risk of death and hospitalisation by age group, socioeconomic status, and vaccination coverage.

Vaccination coverage is higher in the wealthier South and Southeast. Men, people of colour, and low-income groups were more likely to be only partially vaccinated due to missing or delaying a second dose. Vaccination started prematurely for age groups under 50 years and may have hindered uptake of older age groups. Vaccination coverage was associated with a lower risk of death, especially in older age groups (OR: 10.5–34.8, 95% CI: (10.2, 35.9)). Risk of hospitalisation was greater in areas with higher vaccination rates due to higher access to care and reporting.

Vaccination inequality persists between states, age and demographic groups despite increasing uptake. The association between hospitalisation rates and vaccination is attributed to preferential delivery to areas of greater transmission and access to healthcare.

## Introduction

Brazil has experienced the third highest number of confirmed COVID-19 deaths globally. Despite having a universal healthcare system that is experienced with managing public health emergencies^1^, Brazil was unable to contain COVID-19^2^ and intensive care unit (ICU) beds consistently exceeded capacity in many states^3^. This was complicated by the emergence of Gamma (P.1) and spread of Omicron (B.1.1.529) variants^4^. The COVID-19 vaccination campaign was launched across Brazil on January 17, 2021 and initially prioritised healthcare workers, individuals aged 90 years or older, indigenous communities, and institutionalised individuals. Four types of vaccines have been offered to date. However only Coronavac (Sinovac, China) and Covishield AZD1222 (Oxford-AstraZeneca, UK) vaccines were offered during the early phase of the campaign. Vaccine shortages and delays have caused unanticipated interruptions in the administration of doses after the first dose^5^.

During the campaign, dose allocation was determined by the total population of each state without adjusting for demographic differences. Given the increasing risk of SARS-CoV-2 infection and death with age^6^, characterising vaccine uptake across age groups is critical for reducing inequalities and optimizing the distribution of limited vaccine doses. Little is known about how the vaccination programme impacts age-specific hospitalisations and COVID-19-related death rates^7^. To optimise vaccine distribution, further investigation is needed to understand the age-specific reduction in the risk of severe COVID-19 and death, especially among those most vulnerable.

We investigated inequalities in vaccination coverage across Brazil in 2021 and assessed how potential driving factors, such as age, sex, race, and socioeconomic status, influenced coverage. We found several states delayed full vaccination for older populations to prioritise allocation for younger individuals. Among the population that was vaccinated, we estimated the proportion of individuals that did not take the second dose, showing large variation with income, race, sex, and across states. We also estimated the effect of vaccination coverage on the risk of hospitalisation and death from COVID-19 and its associations with socioeconomic conditions. We further assessed the effects of vaccination coverage on death across age groups.

## Methods

### Data sources

#### Vaccination information

Patient level vaccination data notified between 17 January and 6 September, 2021 were retrieved from the Brazilian Ministry of Health’s national immunization system, Sistema de Informação do Programa Nacional de Imunizações (SI-PNI). For every individual that received a dose, we retrieved their age, sex, municipality and state of residence, vaccination facility, dose number (e.g. first or second), and the vaccine type. We did not consider the booster dose campaign in our study, which started on September 15, 2021 with individuals of age 70 years and older.

#### Cases of severe acute respiratory illness

We retrieved data on hospitalisations and deaths in patients with severe acute respiratory illness (SARI) between 1 March, 2020 and 6 September, 2021, from the SRAG (Vigilância de Síndrome Respiratória Aguda Grave) database, which is curated by the Ministry of Health. Patient information on age, sex, and municipality of residence were included. All SARI cases and deaths were notified in the SRAG database regardless of hospitalisation. SARI can be caused by SARS-CoV-2 and is defined by the Ministry of Health as influenza-like illness plus one of the following: dyspnoea, persistent chest pain or hypoxia. We included both confirmed COVID-19 cases and SARI cases with unknown etiology, as those were likely related to COVID-19 but not lab-confirmed due to low rates of COVID-19 testing in Brazil and socioeconomic bias in testing^82^. All SARI cases confirmed to be caused by other respiratory viruses were excluded from the analysis. From the remaining cases, we filtered for (1) all hospitalisations and, (2) all deaths (e.g., hospitalised deaths and other).

#### Socioeconomic conditions and population size

Measurements of socioeconomic indicators at the municipality level were retrieved from the latest population census (2010) compiled by the Brazilian Institute of Geography and Statistics^9^. We selected indicators based on their relevance to the social determinants of health, such as household income per capita, proportion of residents with only a primary education or lower, and unemployment rates. Projected distributions of age and sex for the population in 2020 were obtained for each municipality and state^10^.

### Data analysis

#### Age-group specific vaccination coverage

We estimated the proportion of the population vaccinated with at least one dose using the SI-PNI data and the projected population in each municipality as a whole, and for each age group within that municipality: under 20, 20–29, …, 70–79, and 80 years and above^10^. We used the population estimates to account for the excess deaths caused by COVID-19.

To compare the levels of first dose coverage between older and younger populations, we defined a *campaign optimality index* (COI). The COI for age group A at threshold T is denoted as *η*(*A, T*). The COI measures the proportion of the subsequent (older) age group at the time when the age group *A* first reached its threshold coverage of *T*. To compute the optimality index for threshold *T* and age group *A* = [*a*_min_, *a*_max_] (which includes individuals aged between *a*_min_ and *a*_max_, we calculated the date when the coverage in that age group reached the threshold *T* and then defined *η*(*A, T*) as the coverage of the subsequent age group *A*_older_ = [*a*_min_ + 10, *a*_max_ + 10].

If vaccination was only offered to an age group *A* when everyone in the subsequent age group was vaccinated, then the COI would be 1. Smaller values of the COI for a particular age group indicates that fewer people in the older age group were vaccinated. In an ideal scenario where the vaccination campaign for younger groups only begins after older groups achieve high coverage, the coverage of the older group *A*_older_ should be close to 100% when the coverage of the younger group reaches *T*, in which case *η*(*A, T*) = 1. The campaign optimality index was computed for all age groups and using thresholds of 25%, 50% and 75%.

#### Population that missed their second dose

We estimated the proportion of the population that delayed the second dose, i.e. the proportion did not receive their second dose within the suggested timeframe (21–28 day interval for Coronavac, and 90 days for all other approved 2-dose COVID-19 vaccines). To estimate the proportion of the population that missed the second dose entirely, we determined (i) the proportion of all first dose vaccinated individuals that did not have a second dose recorded, and (ii) from this, selected only those for which the time interval between the first dose and the date of the last entry in the dataset (September 06, 2021) is higher than the suggested time interval between doses for the corresponding vaccine plus a tolerance of 30 days. In other words, this is the proportion of individuals that did not take the second dose and whose second dose is delayed by at least 30 days. Both proportions were estimated for each age, sex, race and municipality. We excluded the single-dose vaccine of Janssen-Cilag from this analysis.

#### Risk of COVID-19 death by age-group

We used logistic regression to estimate the probability that a SARI patient would die with COVID-19. Due to substantial amount of missing vaccination data (54% of all SARI patients with symptoms onset before September 6th, reaching as high as 87% before March 1st), and uncertainty about the reason the data was missing, we used age group and sex specific vaccination coverage of the individual’s municipality of residence as a proxy. For each SARI patient, we used their age group, the socioeconomic conditions, and vaccination coverage of their municipality of residence. We also included a binary indicator for whether the SARI case was hospitalised before the start of the vaccination campaign for their age group. See the SI for further details on the model used.

#### Risk of COVID-19 hospitalisation by age group

We used quasi-Poisson regression (GLM with a log-link function) to model the number of SARI hospitalisations in each state adjusting for age group, whether the hospitalisations occurred before or after the start of the vaccination campaign (for each age group and state), and each state’s average age group specific vaccination coverage and socioeconomic factors such as the level of unemployment, education, and per capita income. Given data availability, we used the aggregated values for socioeconomic status across each state as a proxy for the individual’s values. See the SI for further details on the model used.

#### Age distribution of death by vaccination coverage

To assess the potential protective effect of vaccination coverage by age, we looked at the distribution of ages among COVID-19 deaths in the SARI database. We modelled the ages as a sample from a multinomial distribution with parameter *θ* = (*θ*_1_, …, *θ*_*n*_) where *θ*_*j*_ is the probability of a death belonging to the *j*th age group. The posterior distribution of *θ* is a Dirichlet distribution with parameters *α* = (1 + *D*_1_, …, 1 + *D*_*n*_), where *D*_*j*_ is the number of deaths observed in age group *j*, assuming a non-informative Dirichlet prior distribution with parameters (1, …, 1). Sampling from the posterior distribution provided the marginal credible intervals for the proportion of COVID-19 deaths in each age group at four points in the epidemic: Before January 17, January 17 to March 16, March 17 to May 16, and May 17 to September 16, 2021.

## Results

### Geographical inequalities in vaccination coverage

Between January 17 and September 6, 2021, 133.4 million people were recorded with a first dose, corresponding to 84.4% of the population above 18 years of age. We determined the proportion of the population vaccinated with at least one dose in each municipality and mapped the distribution of coverage across states (Figure 1A). Similar trends can be observed for the proportion of the population that is fully vaccinated (Figure S1). At the national level, older age groups have higher vaccination coverage, which decreases with age (Figure 1B). A greater proportion of women were vaccinated than men; this difference is more evident among younger age groups below the age of 50 years. Figure 1C shows that there is a clear regional divide in vaccination coverage, with the South and Southeast states having the highest coverage, and the North and Northeast states having the lowest coverage, although there is a substantial amount of variability between the municipalities within each state.

**Figure 1:**
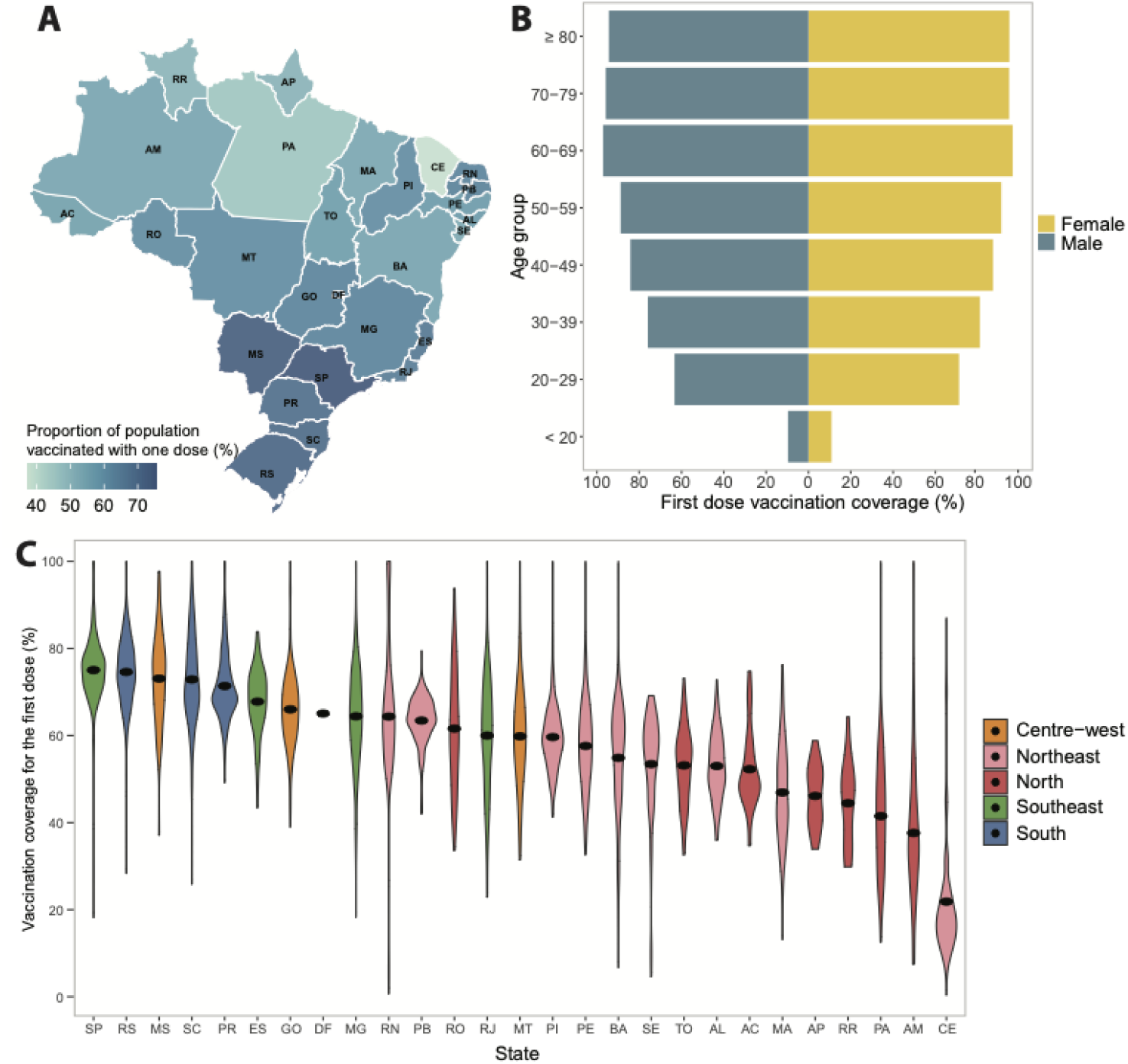
Inequalities in COVID-19 vaccination coverage across Brazil from January 17–September 6, 2021. (A) Percentage of population vaccinated with at least one dose across states as of September 6, 2021. (B) Distribution of vaccination uptake for the first dose between men and women by age group. (C) Vaccination coverage across municipalities (black dot represents the mean coverage).

### Premature advancement of the vaccination campaign towards younger groups

Figure 2A shows the proportion of the population vaccinated with a first dose in Brazil, by age group and sex over time. Due to uncertainty in total population size, coverage exceeds 100% for some groups. Similarly, coverages smaller than 100% may not imply the remaining population is not immunized. At the start of the campaign, both healthcare workers and older populations were prioritised for vaccination, hence delayed vaccination for younger groups. The vaccination coverage by sex shows the coverage for men remained consistently lower than women in all age groups over time.

**Figure 2:**
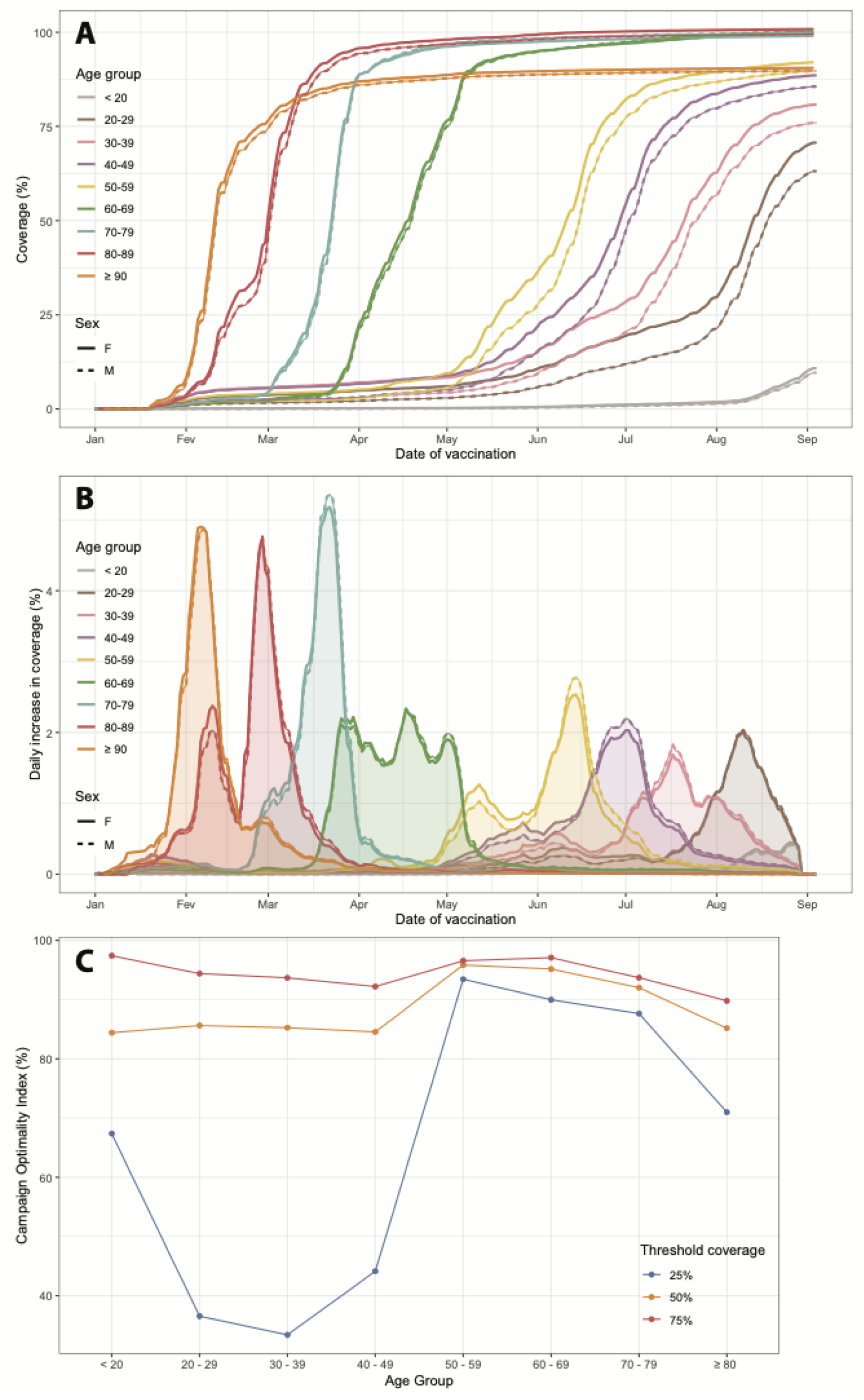
COVID-19 vaccination uptake over time among age groups. (A) Vaccination coverage for the first dose over time. (B) Daily change in vaccination coverage by age group and sex. (C) Campaign optimality index (COI) by age group using threshold vaccination coverage of 25%, 50% and 75%. This index could not be calculated for the age group 20–29 using the threshold 75% because this group had not reached a coverage of 75% by September 6th 2021.

We observed an unanticipated increase in coverage among younger age groups before older groups were fully vaccinated (Figure 2B). This pattern is accentuated in populations below 60 years of age. Using the Campaign Optimality Index (COI), we defined the coverage of older groups when the coverage of younger groups reached a threshold coverage of 25%, 50% and 75%. Larger values indicate a more staggered distribution indicating stronger prioritisation towards older age groups, while lower values indicate that age groups are being vaccinated together (i.e. weaker prioritisation). In Figure 2C, COI is higher for age groups above 50 years old, and drops substantially for younger groups. The difference between age groups is greatest for the threshold of 25% and decreases with higher thresholds, with COI being above 80% for all age groups for the threshold of 75%. Even though the vaccination campaign for younger groups started prematurely, the coverage of younger groups crossed the threshold of 75% when coverage for older groups was higher than 80%.

### Probability of missing the second dose varies by sex, race, and income

By September 6, 2021, 11.9% and 10.2% of the individuals vaccinated with the first dose had their second dose, respectively, delayed and missed. Only 14.3% of individuals that did not take the second dose after 30 days past the recommended date ended up taking a delayed second dose. Among the individuals that were vaccinated with a first dose of Coronavac, 8.78% (95% CI 8.77%–8.80%) missed their second dose. This is low compared to the 12.27% of individuals that missed the second dose (95% CI 12.25%–12.29%) of other vaccines (Covishield and Pfizer), where the optimal interval between the first and second dose is 90 days.

The probability of missing the second dose by race and sex is shown in Figure 3A. Men were more likely to miss (Relative Risk(RR) 1.065, 95% CI 1.063–1.067) the second dose than women. Indigenous Brazilians were also 2.64 (95% CI 2.62–2.66) times more likely to miss the second dose than White Brazilians, followed by East Asian Brazilians (RR 1.68, 95% CI 1.68–1.69), Pardos (RR 1.65, 95% CI 1.65–1.65) and Black Brazilians (RR 1.51, 95% CI 1.50–1.51).

**Figure 3:**
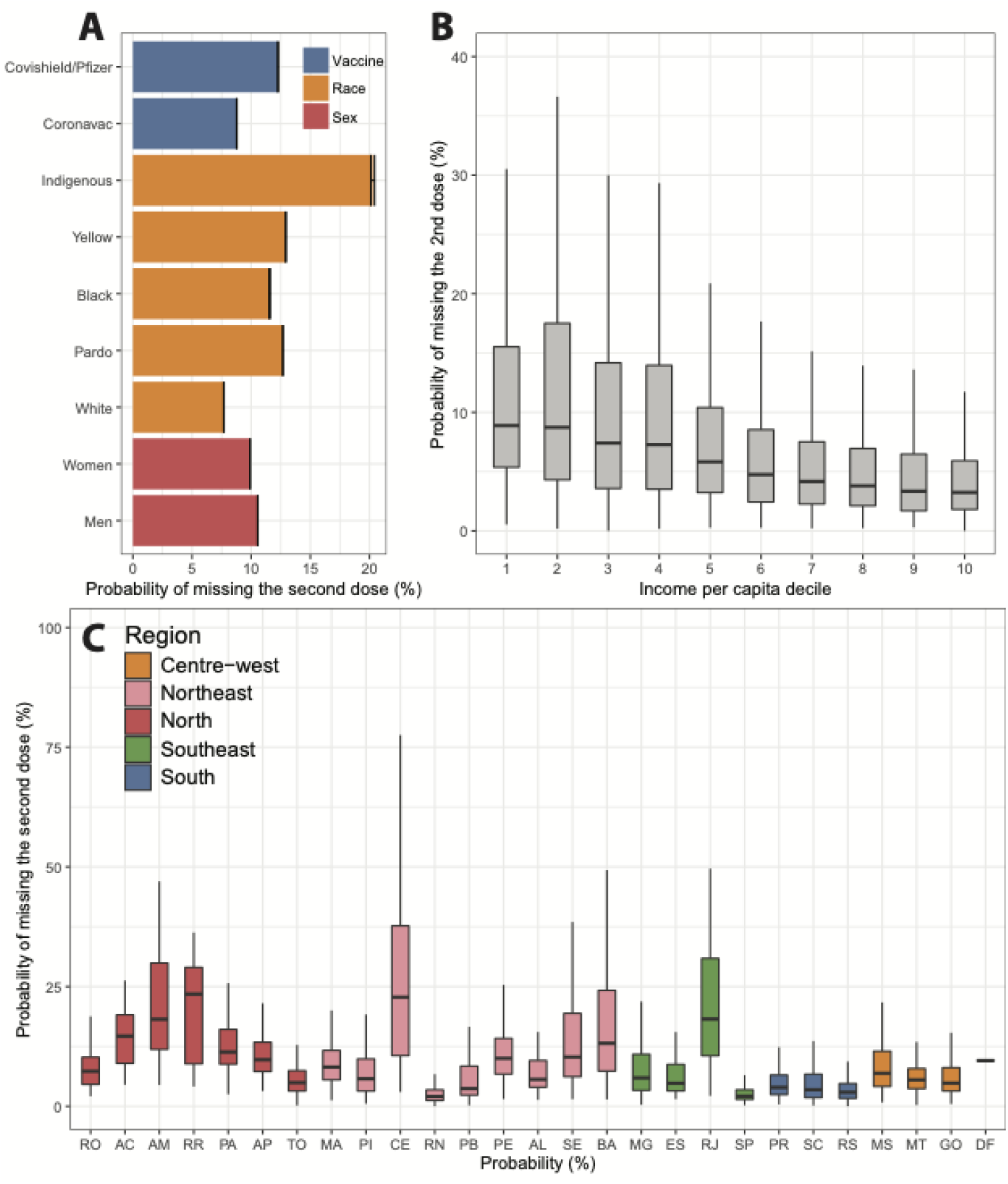
Probability of missing the second dose of the COVID-19 vaccine. (A) Probability of missing the second dose based on three factors: vaccine type, race and sex. (B) Probability of having a second dose missed calculated at municipality level in terms of the income per capita decile. (C) Probability of missing the second dose calculated for each municipality and grouped by state.

The probability of missing the second dose varies with income per capita (Figure 3B). Individuals in municipalities with a lower income per capita are more likely to miss their second dose. The median income per capita for municipalities with a 2 % probability of skipping the second dose is R$546,80 (IQR R$421,60–R$650,50). This decreases to R$270,40 (IQR R$212,60–R$420,80) when the probability of skipping is more than 10%. Figure 3C shows the variation in the probability of missing the second dose by state.

### Vaccination decreases age-adjusted risk of death

Vaccination decreases the risk of COVID-19 death after adjusting for age group and socioeconomic conditions. Using 20 years of age and under as a baseline, the risk of COVID-19 death increased with age. For instance, the odds of death are at least ten times greater for someone above the age of 50 years (OR: 10.5–34.8, 95% CI: 10.2, 35.9) than an individual under 20 years of age. Patients living in municipalities with higher income per capita (OR: 0.919, 95% CI: 0.917, 0.920) and lower education attainment (OR: 0.934, 95% CI: 0.932, 0.936), respectively, had reduced risk of death. On the contrary, patients from municipalities with higher unemployment rates were slightly more likely to be at risk of death (OR: 1.009, 95% CI: 1.007, 1.010). When the vaccination campaign was accounted for, patients were at a lower risk of death after the start of the vaccination campaign (OR: 0.834, 95% CI: 0.832, 0.837); patients living in municipalities with higher vaccination coverage also had lower risk (OR: 0.996, 95% CI: 0.994, 0.997). Our sensitivity analysis at the individual level which includes only patients with a recorded vaccination status, showed that patients were at a lower risk of death after being vaccinated with at least one dose. While similar patterns for age groups were observed for hospitalisations, higher unemployment was associated with lower risk of hospitalisation. Detailed information on model outputs can be found in Table S1 and Table S2.

To verify the relationship between vaccination coverage and risk of death by age groups, we plotted the probability of a patient dying from COVID-19 being in a particular age group prior to January 17, 2021 and after the start of the vaccination campaign (including and after January 17, 2021) (Figure 4A). There is a higher probability that those dying before the start of the vaccination campaign were from an older age group. After vaccination began, those dying were more likely to be from age groups with ages below 60 years. This can be attributed to vaccination, where the proportion of vaccination is much higher among people in older age groups due to earlier uptake (Figure 4B). However, individuals under 20 years of age showed a reverse trend, where the proportion of deaths was higher prior to the vaccination campaign.

**Figure 4:**
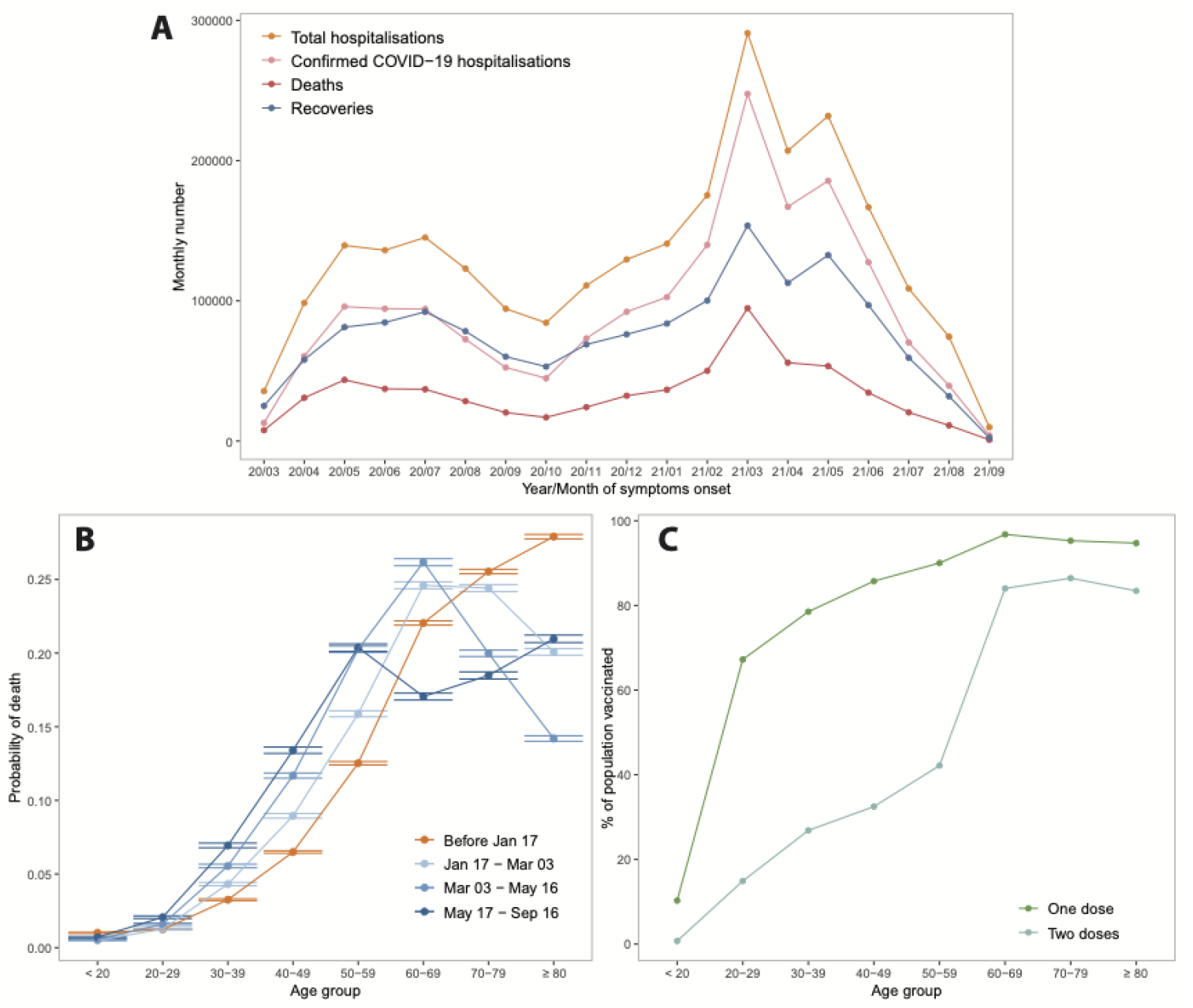
Risk of SARI death and vaccination coverage by age distribution. (A) Distribution of SARI hospitalisations over time (March 2020–September 2021). (B) The distribution of age among SARI-linked deaths at four points in time indicates that as the epidemic progressed SARI deaths were more likely to be younger individuals. (C) Proportion of population vaccinated with at least one dose by age group during study period.

## Discussion

Inequalities exist in COVID-19 vaccination coverage, especially across states and different age, sex, and socioeconomic groups. Among patients diagnosed with COVID-19, the riskof hospitalisation and death are significantly greater among older individuals and those with lower socioeconomic status. Increasing vaccination reduces the risk of death, especially among older age groups. Vaccine uptake is greater in areas where hospitalisation rates are high, suggesting better access to healthcare and improved reporting of infections.

Variation in coverage between population groups was mainly driven by the unequal distribution of vaccines across states. States from the North and Northeast regions, such as Amazonas, Amapá, and Ceará, were prioritised for vaccination in January 2021 due to high transmission rates of the P.1 (Gamma) variant, which was first detected in Manaus (the capital of Amazonas) in January 2021^11^. Despite this, lower overall coverage for the first dose was observed in the North and Northeast regions of Brazil. These regions are economically less developed and have experienced scare hospital resources during the pandemic^12^. Delays in notification of vaccinations had an important impact on coverage between states. Even though we determined a 37.5% first-dose coverage for the northeastern state of Ceará, official statistics obtained directly from the Brazilian state secretariats on September 6, 2021 found this was 59.2% (G1 Globo, 2022). The low COI obtained with a threshold of 25% for age groups younger than 50 shows the vaccination campaign started prematurely for these groups, which may have delayed the vaccination of older groups. It is worth noting that the proposed COI only considers age as a priority criterion and does not include other criteria such as comorbidities or employment (e.g. healthcare worker).

Inequalities in uptake are evident between the sexes. Women were more likely to have received both doses than men. This may be owing to the early stages of the vaccination campaign, when healthcare workers were prioritised: women make up a significant proportion of healthcare workers in Brazil, accounting for 62.5% of all vaccinated healthcare workers, which corresponds to 10.6% of all vaccinated individuals in our dataset. Moreover, women seek out healthcare more frequently than men. Based on a survey study with over 170,000 responses, men showed greater hesitancy towards COVID-19 vaccines, indicating the main reason being fear of severe clinical reactions^13^. We also found a longer delay for taking the second dose in municipalities with lower income per capita, which is aligned with previous findings that low incomes were associated with higher rates of vaccine hesitancy during the pandemic^13^. This has implications for death risk, as our findings show that SARI patients from poor municipalities are more likely to die from COVID-19 than those from wealthier municipalities.

Vaccination reduces the risk of death. The positive association between vaccination and hospitalisation appears to be driven by greater accessibility of both in wealthier areas. To further investigate whether vaccination can prevent COVID-19 related deaths for each age group, we considered the age distribution of those that died, across multiple periods of time. We observed a reduction in the proportion of mortality in older age groups once vaccination became available, since it was the elderly that were prioritised for vaccination. This trend was the least evident with young people under the age of 20. This was likely owing to the delayed vaccination of young people until July 2021. Due to a lack of complete population-level data, it may be premature to surmise the true protective effects of vaccination coverage on younger age groups. While the overall risk of death is lower for young people, COVID-19 still poses a threat, regardless of pre-existing medical conditions^14,15^.

Our estimates of vaccination coverage by state and age-group are limited by the accuracy of the projected population estimates. We were unable to retrieve the vaccination status of individual SARI patients and thus had to rely on overall vaccination coverage for each age group; this may have introduced discrepancies in the modelled relationship between vaccination coverage and the risk of death. Furthermore, human error affects the accuracy of vaccination records (e.g. date of vaccination could be replaced by the date when the information was entered into the system). Moreover, the lack of healthcare resources in some areas (e.g. Ceará) may have led to lower levels of case notification and notification delay. Self-identification of race, may be prone to bias, with wealthier Black Brazilian more inclined to self-identify and contribute race information than poorer Black Brazilians^16^.

Despite some vaccine hesitancy among certain demographic groups, the overall intention to be vaccinated against COVID-19 is high^17,13^. Vaccine hesitancy is a complex issue; a survey from late 2020 found that when certain countries of origin for the vaccine were mentioned (e.g. China and Brazil), the willingness to vaccinate among Brazilians decreased. However, our study shows that individuals that were vaccinated with the first dose of Coronavac were less likely to skip the second dose when compared with individuals that were vaccinated with other vaccines, such Covishield or Pfizer. This may be partially attributed to the fact that the optimal interval between the first and second doses for most vaccines is 90 days, while the interval for Coronavac is only 21 to 28 days^18^. The disparities in vaccination coverage may have amplified the inequalities in risk of death during the epidemic in Brazil, as the Gamma variant is associated with an increased infection fatality rate^19^.

The third booster dose has been controversial as parts of the population were still awaiting their second dose. Vaccination has a protective effect on populations^20^ but the details of how this is communicated to the general public are complex^20^. Our findings on inequalities in coverage and on demographic groups that are more willing to miss the second dose should be considered when designing media campaigns and interventions to increase vaccine access and uptake.

## Data Availability

All data produced in the present study are available upon reasonable request to the authors.

https://github.com/sabrinalyli/BrazilCOVID19Vaccination

## Funding

SLL was supported by the Oxford Martin Programme on Pandemic Genomics and the Canadian Social Sciences and Humanities Research Council (SSHRC) Doctoral Fellowship. CAPJ is supported by FAPESP (2019/21858-0 and 2022/15985-1). AEZ was supported by the Oxford Martin School Programme on Pandemic Genomics. CAPJ and VHN were supported by Coordenação de Aperfeiçoamento de Pessoal de Nível Superior – Brasil (CAPES) – Finance Code 001. VHN is supported by the Brazilian National Council for Scientific and Technological Development (CNPq: 304714/2018-6). ECS is supported by a Medical Research Council-São Paulo Research Foundation (FAPESP) CADDE partnership award (MR/S0195/1 and FAPESP 18/14389-0) (http://caddecentre.org/).

## Ethics

This project was approved by the Brazilian National Research Ethics Committee (CONEP CAAE-30178220.3.1001.0068).

## Author Contributions

- Sabrina L. Li - Conceptualization, Data Curation, Formal Analysis, Investigation, Methodology, Software, Validation, Visualization, Writing – Original Draft Preparation, Writing – Review & Editing
- Carlos A. Prete Jr - Conceptualization, Data Curation, Formal Analysis, Investigation, Methodology, Software, Validation, Visualization, Writing – Original Draft Preparation, Writing – Review & Editing
- Alexander E. Zarebski - Conceptualization, Formal Analysis, Methodology, Software, Validation, Writing – Original Draft Preparation, Writing – Review & Editing
- Andreza A de Souza Santos – Investigation, Validation, Writing – Review & Editing
- Ester C Sabino - Investigation, Validation, Writing – Review & Editing
- Vitor H Nascimento - Formal Analysis, Validation, Writing – Review & Editing
- Chieh-Hsi Wu - Formal Analysis, Validation, Writing – Review & Editing, Supervision
- Jane P Messina - Conceputalization, Formal Analysis, Writing – Review & Editing, Supervision

## Notes

### Competing Interest Statement

The authors have declared no competing interest.

### Author Declarations

This project was approved by the Brazilian National Research Ethics Committee (CONEP CAAE-30178220.3.1001.0068). This study used only openly available data that were originally located at https://opendatasus.saude.gov.br/dataset/srag-2020 https://opendatasus.saude.gov.br/dataset/srag-2021-a-2023 https://opendatasus.saude.gov.br/dataset/covid-19-vacinacao

