## Supplementary Material for "The Brazilian COVID-19 vaccination campaign: A modelling analysis of socio-demographic factors on uptake"

[1]School of Geography and the Environment, University of Oxford, Oxford, UK
[2]School of Geography, University of Nottingham, Nottingham, UK
[3]Department of Electronic Systems Engineering, University of São Paulo, São Paulo, Brazil
[4]Department of Biology, University of Oxford, Oxford, UK
[5]School of Mathematics and Statistics, University of Melbourne, Melbourne, Australia
[6]Oxford School of Global and Area Studies, Latin American Centre, University of Oxford, Oxford, UK
[7]Departamento de Molestias Infecciosas e Parasitarias & Instituto de Medicina Tropical da Faculdade de Medicina da Universidade de São Paulo, São Paulo, Brazil
[8]Mathematical Sciences, University of Southampton, Southampton, UK
[9]Oxford School of Global and Area Studies, University of Oxford, Oxford, UK
[=]These authors contributed equally

#### Risk of COVID-19 death by age-group

The equation describing the log-odds of death among SARI patients is expressed as

$$\begin{aligned}\text{logit}(p_i) = \beta_0 &+ \beta_1 \cdot (\text{Age group of } i) \\ &+ \beta_2 \cdot (\text{Vaccination coverage in age group of } i) \\ &+ \beta_3 \cdot (\text{Campaign status for } i) \\ &+ \beta_4 \cdot (\text{Unemployment for age group of } i) \\ &+ \beta_5 \cdot (\text{Low education attainment for age group of } i) + \epsilon_i,\end{aligned} \tag{1}$$

where $p_i = \mathbb{P}(Y_i = 1)$ is the log-odds of death where $Y_i$ is an indicator variable for whether the $i$th SARI patient's outcome: dead = 1, and alive = 0. The $beta_j$ are the coefficients of the model and $\epsilon_i$ is the error term. For reference, the logit function is defined as $\text{logit}(p) = \log(p/(1-p))$.

As a form of sensitivity analysis, we fit the model at the individual level by including only patients that had a recorded vaccination status (e.g. vaccinated with at least one dose, or no vaccination). Here we included vaccination status as a binary indicator in the model to replace the age-group specific vaccination coverage. While this avoids the potential for an ecological fallacy (i.e. assuming population level vaccination levels are indicative of the individuals represented in the data), we cannot rule out the possibility that data may not be missing at random, which may introduce biases in the estimates.

The resulting parameter estimates from this model and their confidence intervals are given in Table S1.

| Variable | Coefficient (95% CI) | Significance |
|---|---|---|
| Intercept | -3.60 (-3.63, -3.57) | * |
| Age group#: 20-29 years old | 1.17 (1.14, 1.21) | * |
| Age group: 30-39 years old | 1.51 (1.14, 1.21) | * |
| Age group: 40-49 years old | 1.91 (1.88, 1.94) | * |
| Age group: 50-59 years old | 2.35 (2.32, 2.37) | * |
| Age group: 60-69 years old | 2.87 (2.84, 2.90) | * |
| Age group: 70-79 years old | 3.21 (3.18, 3.24) | * |
| Age group: 80 years and older | 3.55 (3.52, 3.58) | * |
| Age-group specific coverage | -0.00414 (-0.00568, -0.00261) | * |
| Age-group specific campaign (yes) | -0.182 (-0.186, -0.178) | * |
| Unemployed | 0.0241 (0.0221, 0.0261) | * |
| Primary education or lower | -0.0737 (-0.0758, -0.0716) | * |
| Income per capita | -0.0579 (-0.0602,-0.0556) | * |

### Baseline age group for comparison is under 20 years old
* statistically significant $p$-value $< 0.05$

**Table S1:** Logistic regression model analysis — death risk of SARI patient given age group, socioeconomic status, age group-specific vaccination coverage, and indication of age-specific vaccination campaign.

#### Risk of COVID-19 hospitalisation by age group

The equation describing the expected number of hospitalisations in this model is

$$\begin{aligned}\log(\mathbb{E}[Y_i]) = \beta_0 &+ \beta_1 \cdot (\text{Age group of } i)\\ &+ \beta_2 \cdot (\text{Vaccination coverage in age group of } i)\\ &+ \beta_3 \cdot (\text{Campaign status for } i)\\ &+ \beta_4 \cdot (\text{Unemployment for age group of } i)\\ &+ \beta_5 \cdot (\text{Low education attainment for age group of } i)\\ &+ \beta_6 \cdot (\text{Income percapita for age group of } i) + \epsilon_i,\end{aligned} \tag{2}$$

with the variance $\text{Var}(Y_i) = \phi(Y_i)$ where $\phi$ is the overdispersion parameter and $\epsilon_i$ is the error term.

The resulting parameter estimates from this model and their confidence intervals are given in Table S2.

| Variable | Coefficient (95% CI) | Significance |
|---|---|---|
| Intercept | -6.67 (-7.45, -5.89) | * |
| Age group#: 20-29 years old | -0.0427(-0.161, -0.246) | |
| Age group: 30-39 years old | 0.884 (0.721, 1.05) | * |
| Age group: 40-49 years old | 1.39 (1.24, 1.54) | * |
| Age group: 50-59 years old | 1.84 (1.69, 1.98) | * |
| Age group: 60-69 years old | 2.19 (2.04, 2.34) | * |
| Age group: 70-79 years old | 2.62 (2.47, 2.77) | * |
| Age group 80 years and older | 3.16 (3.02, 3.30) | * |
| Age-group specific coverage | 0.00875(0.000612, 0.0169) | * |
| Age-group specific campaign (yes) | 0.0533 (-0.000682, 0.115) | |
| Unemployed | -3.98 (-5.65, -2.31) | * |
| Primary education or lower | 1.94 (0.502, 3.37) | * |
| Income per capita | -0.000363 (0.0000902,0.0000982) | * |

### Baseline age group for comparison is under 20 years old
* statistically significant $p$-value $< 0.05$

**Table S2:** Quasi-Poisson regression model analysis — hospitalisation risk given age group, socioeconomic status, age group-specific vaccination coverage, and indication of age-specific vaccination campaign

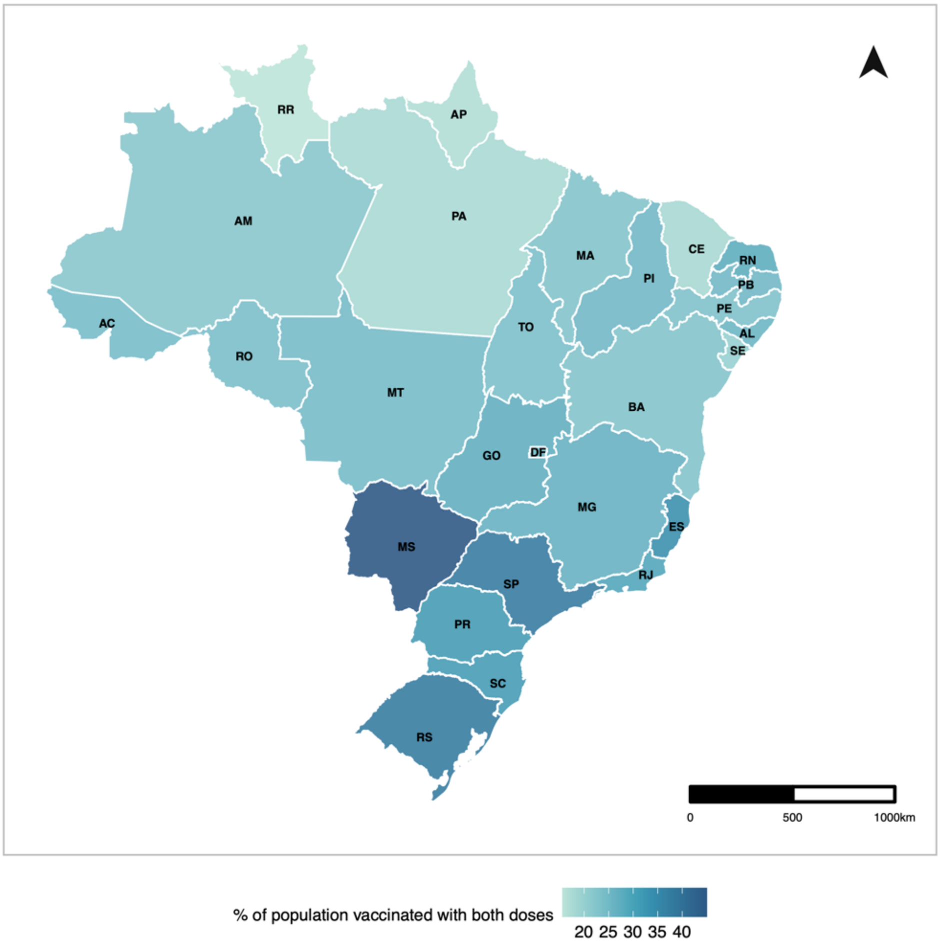


**Figure S1:** Proportion of population fully vaccinated.
